## Supplemental file 1 for "The impacts of social restrictions during the COVID-19 pandemic on the physical activity levels of over 50-year olds: the CHARIOT COVID-19 Rapid Response (CCRR) cohort study"

**Supplementary File 1**

Variables Extracted from Survey

| **Physical Characteristics** | **Lifestyle Characteristics** | **Social Isolation Characteristics** |
| --- | --- | --- |
| Age | Alcohol drinker (yes/ no) | Number of people in household |
| Sex | Smoking status (yes/ no) | Relationship status (in a relationship/ single) |
| Height | Six variables from pre-restriction IPAQ results | Shielding status (yes/ no) |
| Weight | Six variables from IPAQ results for the week prior to the survey | Loneliness frequency (never/ rarely/ sometimes/ often) |
| Ethnicity |  |  |
| Underlying conditions (present/ absent) |  |  |
