## Supplemental file 2 for "The impacts of social restrictions during the COVID-19 pandemic on the physical activity levels of over 50-year olds: the CHARIOT COVID-19 Rapid Response (CCRR) cohort study"

**Supplementary File 2**

S2.1 Causal diagrams for loneliness

Model 1

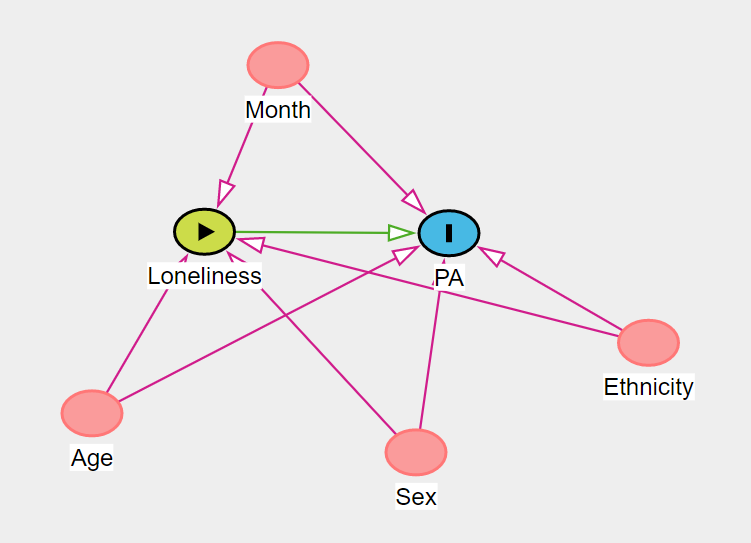

Model 2

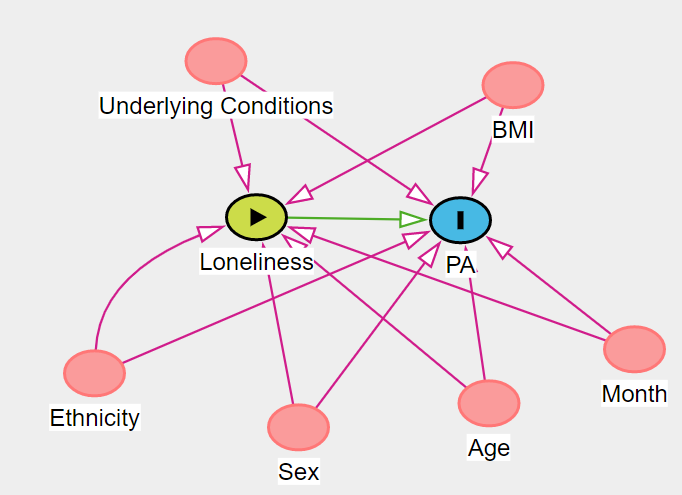

Model 3

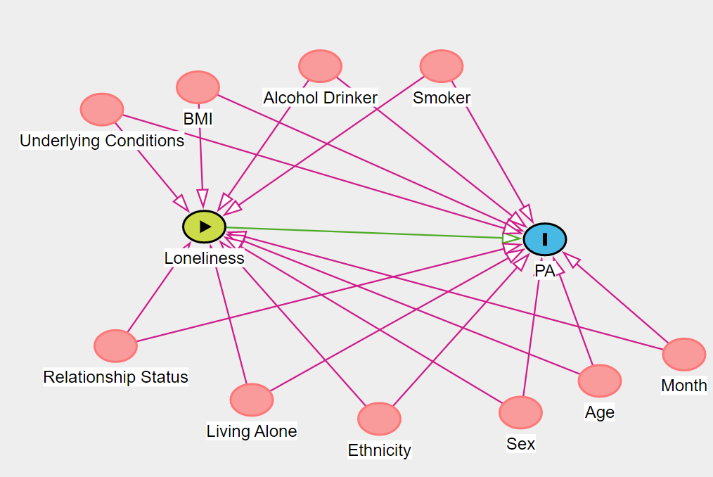

S2.2 Causal diagrams for shielding

Model 1

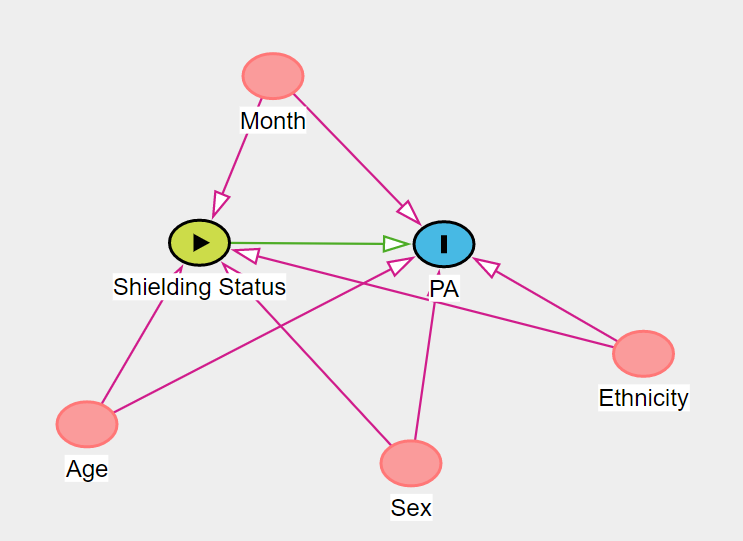

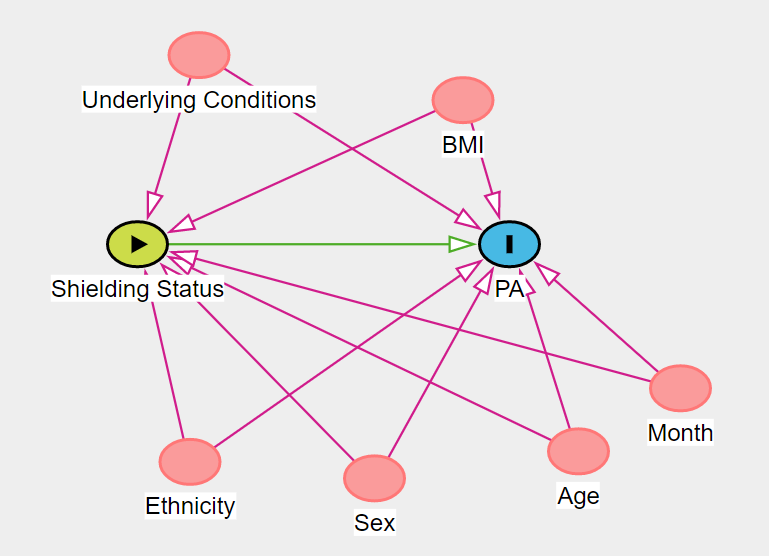
Model 2

Model 3

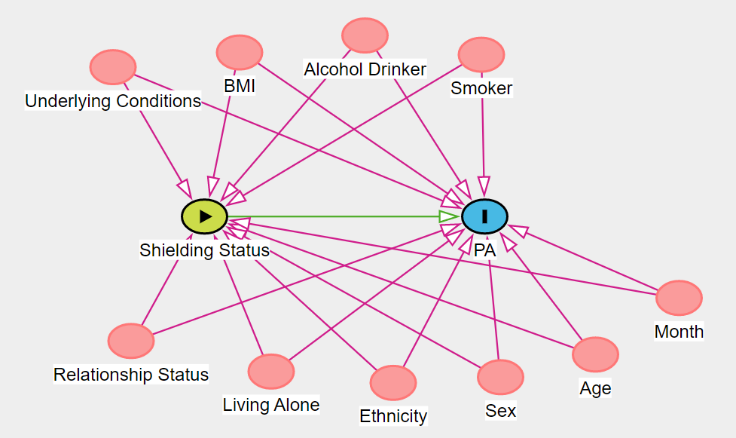
