## Supplementary figures and images for "The impacts of social restrictions during the COVID-19 pandemic on the physical activity levels of over 50-year olds: the CHARIOT COVID-19 Rapid Response (CCRR) cohort study"

### Supplemental file 3

**Supplementary File 3**

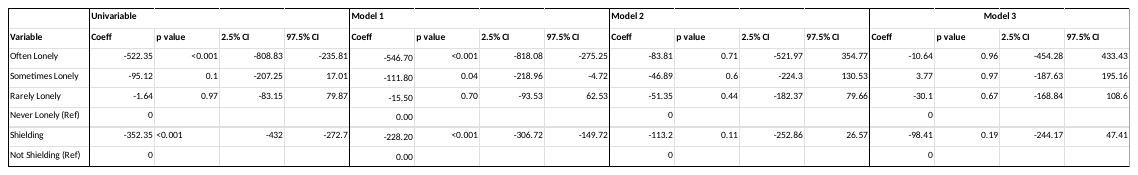
Coefficients of Study Variables
